# Hundreds of severe pediatric COVID-19 infections in Wuhan prior to the lockdown

**DOI:** 10.1101/2020.03.16.20037176

**Authors:** Zhanwei Du, Ciara Nugent, Benjamin John Cowling, Lauren Ancel Meyers

## Abstract

Before January 22, 2020, only one pediatric case of COVID-19 was reported in mainland China^1,2^. However, a retrospective surveillance study^3^ identified six children who had been hospitalized for COVID-19 in one of three central Wuhan hospitals between January 7th and January 15th. Given that Wuhan has over 395 other hospitals, there may have been far more severe pediatric cases than reported.

---

There were six and 43 children out of 336 who tested positive for COVID-19 and influenza, respectively among all pediatric admissions during the 9-day period^3^. By using this ratio in a detailed analysis of influenza surveillance data and COVID-19 epidemic dynamics (see Appendix), we estimate that there were 313 [95% CI: 171-520] children hospitalized for COVID-19 in Wuhan during January 7-15, 2020 (Figure). Under an epidemic doubling time of 7.31 days^4^, we estimate that there were 1105 [95% CI: 592, 1829] cumulative pediatric COVID-19 hospitalizations prior to the January 23rd lockdown, which far surpasses the 425 confirmed cases reported across all age groups, none of which were children under age 15^1^.

**Figure.**
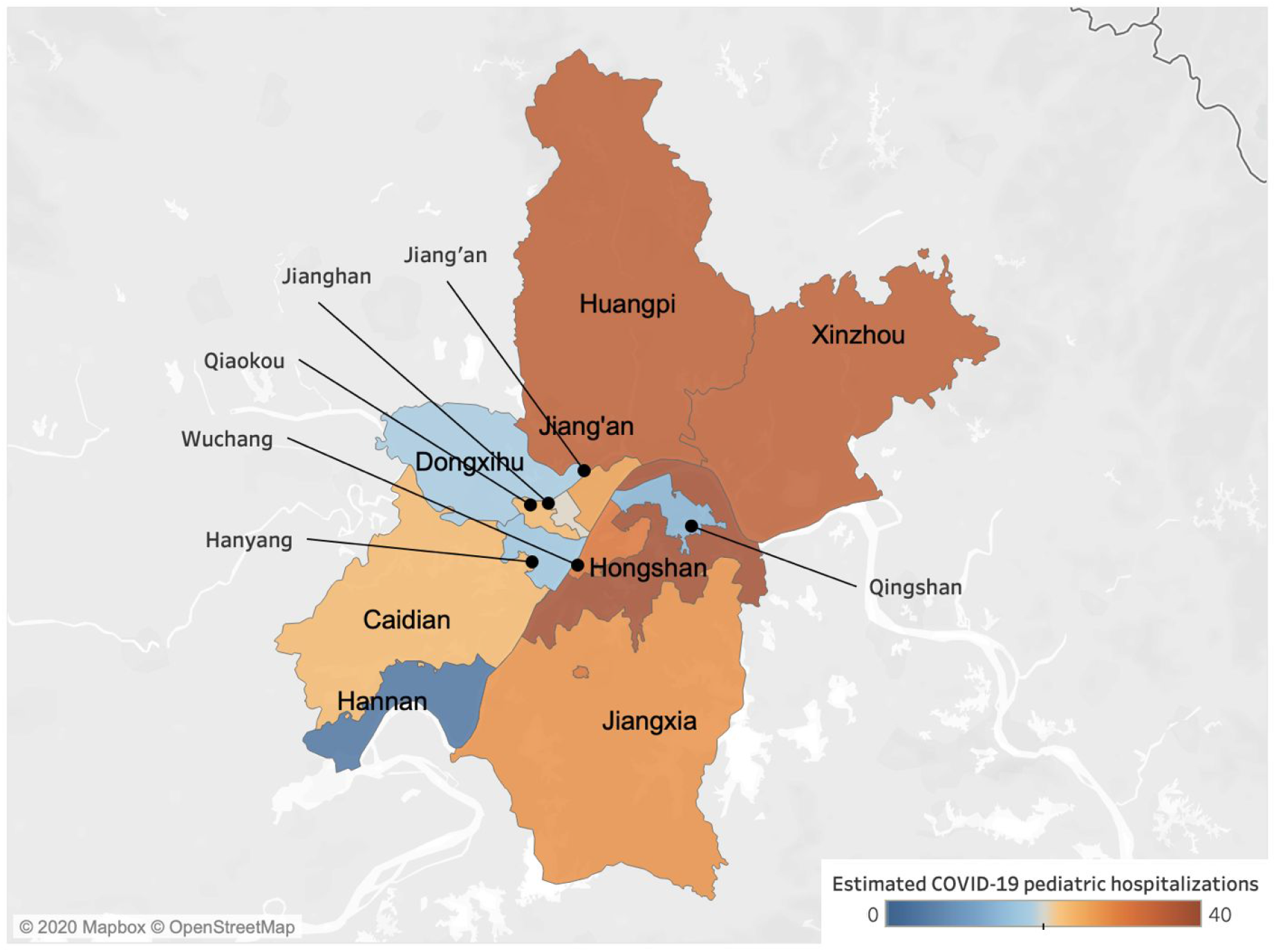
Estimated COVID-19 pediatric hospitalizations (under age 17) in the 13 districts of Wuhan from January 7 to January 15, 2020. A retrospective study identified five hospitalized pediatric cases of COVID-19 from four central districts of Wuhan^3^ and one from the neighboring Huanshi district^3^. We estimate that there were a total of 313 [95% CI: 171-520] severe (hospitalized) cases of COVID-19 in children during that nine-day period across the 13 central districts of Wuhan. Across districts our estimates range from four [95% CI: 0-9] in suburban Hannan to 38 [95% CI: 19-68] in central Hongshan as indicated by shading (Table S1).

Children are strikingly absent from COVID-19 reports and limited data suggest that pediatric infections are overwhelmingly mild^5^. Thus, our estimates for hundreds of severe pediatric cases likely translates to thousands or even tens of thousands of mildly infected children, suggesting that the force of infection from children may be grossly underestimated and the infection fatality rate overestimated from confirmed case counts alone. This highlights the urgent need for more robust surveillance to gauge the true extent and severity of COVID-19 in all ages.

## Data Availability

All available in open access

## Acknowledgments

We acknowledge grant support from NIH (U01 GM087719).

## Author Contributions

Zhanwei Du, Ciara Nugent and Lauren Ancel Meyers: conceived the study, designed statistical methods, conducted analyses, interpreted results, wrote and revised the manuscript. Benjamin J. Cowling: conceived the study, interpreted results, and revised the manuscript.

## Ethics committee approval

Not applicable.

## References

1. Li Q, Guan X, Wu P, et al. Early Transmission Dynamics in Wuhan, China, of Novel Coronavirus–Infected Pneumonia. N Engl J Med 2020; published online Jan 29. DOI:10.1056/NEJMoa2001316.

2. Chan JF-W, Yuan S, Kok K-H, et al. A familial cluster of pneumonia associated with the 2019 novel coronavirus indicating person-to-person transmission: a study of a family cluster. Lancet 2020; 395: 514–23.

3. Liu W, Zhang Q, Chen J, et al. Detection of Covid-19 in Children in Early January 2020 in Wuhan, China. N Engl J Med 2020; published online March 12. DOI:10.1056/NEJMc2003717.

4. Zhanwei Du, Lin Wang, Simon Cauchemez, et al. Risk for Transportation of 2019 Novel Coronavirus Disease from Wuhan to Other Cities in China. Emerging Infectious Disease journal 2020; 26. DOI:10.3201/eid2605.200146.

5. Tang A, Xu W, Shen M, et al. A retrospective study of the clinical characteristics of COVID-19 infection in 26 children. Infectious Diseases (except HIV/AIDS). 2020; published online March 10. DOI:10.1101/2020.03.08.20029710.

